# Step Counts and Physical Activity Trends: A 3-Month Analysis of a Walking Challenge Among 30 Female Participants

**DOI:** 10.1101/2024.10.23.24315952

**Authors:** Olajumoke Esan, Taiwo Esan

## Abstract

The Women Walking Challenge (WWC) aimed to increase physical activity among 30 female participants by implementing a structured three-month program from May 1 to July 31, 2024.This study examined daily step count data using wearable devices and fitness Applications to assess activity patterns, consistency, and goal achievement. The results showed an initial increase in step counts, peaking in June at 4,053,122 steps, followed by a decrease in July. Participants took an average of 13,200 steps per day on weekdays and 9,800 on weekends, demonstrating differences in activity levels. Top performers achieved the 10,000-step-per-day goal on more than 75% of days, while low-activity participants struggled to stay consistent. The clustering analysis divided participants into low, moderate, and high-activity groups, revealing distinct engagement patterns. Although the challenge successfully increased physical activity, maintaining long-term motivation remains difficult. These findings offer insights into how to design future health programs that incorporate strategies such as continuous feedback and tailored interventions to support sustained engagement and physical activity.

## Introduction

Physical activity is a cornerstone of both physical and mental health, offering a multitude of benefits that span the entire lifespan. Regular moderate-intensity activities, such as brisk walking, have been shown to significantly impact overall well-being and reduce health risks. Engaging in at least 150 minutes of moderate-intensity aerobic activity weekly has been associated with a 30% reduction in all-cause mortality and a 20% reduction in cardiovascular-related mortality (Kraus et al., 2019). Beyond physical benefits, increased physical activity levels have been linked to enhanced mental health outcomes, including reductions in anxiety and depression symptoms, improved cognitive function, and an overall boost in quality of life (Naylor et al., 2024).

Walking, as one of the most accessible and convenient forms of exercise, stands out for its affordability, low-impact nature, and suitability for people of all fitness levels and ages. Unlike other forms of exercise that may require specialized equipment or facilities, walking can be easily incorporated into daily routines, making it a particularly viable intervention for increasing physical activity. It is especially valuable for populations facing barriers to more strenuous exercise, such as older adults or individuals with limited mobility (Gupta & Vaqar, 2022). Due to its broad applicability and extensive health benefits, walking has become a focal point in public health campaigns and community-based initiatives aimed at combating physical inactivity.

With the advent of wearable technology and fitness tracking devices, step count analysis has become a popular tool for monitoring and encouraging physical activity. Step challenges, which involve participants setting and striving to meet specific daily or weekly step goals, are now widely adopted in community, workplace, and clinical settings to promote physical activity (Schaller et al., 2024). Research has shown that these structured challenges are effective in increasing physical activity levels and delivering various health benefits, including improved weight management, blood pressure regulation, and mental well-being (Safi et al., 2024; Page et al., 2020).

For example, Chaudhry et al. (2020) conducted a systematic review and meta-analysis, finding that participants with specific step count goals significantly increased their daily steps compared to those with more generalized activity goals. This study also found that participants engaged in step count challenges were more likely to meet physical activity recommendations and maintain their engagement over time, highlighting the motivational impact of goal-oriented walking programs. Similarly, Ryu et al. (2023) examined the effects of a step challenge on adults, reporting that participants boosted their average daily step count by around 2,000 steps after joining the program, reinforcing the effectiveness of step challenges as a tool for increasing physical activity.

Despite these promising results, individual responses to step challenges can vary, influenced by factors such as baseline activity levels, personal motivation, social support, and the way goals are structured (Page et al., 2020). Recognizing these factors is crucial for designing step challenge programs that are both inclusive and effective in meeting diverse needs.

In this context, the Women Walking Challenge (WWC) was developed to engage 30 female participants in a three-month structured walking program. The WWC aims to improve participants’ health and well-being through regular walking while creating a supportive environment that leverages motivation and social support to promote sustained engagement. This initiative aligns with existing research, underscoring that well-designed step challenges can serve as powerful motivators, fostering lasting behavior change and supporting overall health improvement across various populations.

## Purpose and Scope of the Study

The primary goal of this research is to assess the impact of the WWC on participants’ physical activity levels by analyzing daily step counts over three months (May 1 to July 31, 2024). The study’s objectives were to identify walking behavior trends or patterns in activity levels, assess participants’ consistency in meeting step goals, and compare activity levels across months. This analysis will shed light on how structured walking challenges affect physical activity engagement and overall health outcomes. The study conducts detailed visual and statistical analyses, allowing for interactive exploration of the dataset. This will aid in understanding the dynamics of physical activity during the challenge and provide evidence-based recommendations for future health promotion initiatives.

### Significance of Study

This study is significant in a variety of ways. First, it adds to the growing body of evidence on the efficacy of step count challenges as a health promotion tool. Previous research has demonstrated that step challenges can increase physical activity levels, promote weight loss, and improve cardiovascular health (Schaller et al., 2024; Ryu et al., 2023). The current study adds to this evidence by conducting a detailed analysis of participant behavior and activity trends over time.

Furthermore, the study makes practical recommendations for future walking challenges and health programs. Understanding the factors that influence participant engagement and consistency allows health practitioners, community organizers, and policymakers to design more targeted and effective interventions to increase physical activity and improve public health outcomes. This research is thus an important resource for developing strategies that promote healthier lifestyles through structured physical activity programs.

### Motivational Factors in Physical Activity Programs

Motivation is essential for both initiating and maintaining physical activity behaviors. According to the Self-Determination Theory (SDT), people who feel autonomous, competent, and connected to others are more likely to engage in physical activity (Gagné et al., 2022).

Motivational factors like goal setting, social support, and feedback have been shown to improve engagement and adherence in walking challenges (Ryan & Deci, 2020). For example, studies show that when participants receive regular feedback on their progress, participate in group activities, or compete against others, they are more likely to meet their daily targets (Smith et al., 2023).

Goal setting specifically, has emerged as an effective motivator in step count challenges. Setting specific, measurable, and attainable goals can help participants focus and serve as a success benchmark (Chevance et al., 2021). In addition, incentives and rewards have been shown to increase motivation and sustain participation over longer periods (Akinrolie et al., 2024). Social factors like group cohesion and peer support help motivate participants by instilling a sense of accountability and shared purpose (Ryu et al., 2023).

### Gaps in Research and Study Contribution

While previous research has demonstrated the benefits of step count challenges in increasing physical activity, several gaps remain. First, most studies have focused on short-term interventions lasting a few weeks or months, with little research looking into the long-term sustainability of behavior change after step challenges (Schaller et al., 2024). Understanding how participants maintain or return to previous activity levels following the completion of a step challenge is critical for designing interventions with long-term effects.

In addition, more research is needed to understand how individual differences, such as age, fitness level, and baseline activity, affect the effectiveness of step count challenges. While some studies have identified personal factors that predict success in step challenges, more comprehensive analyses are needed to investigate how these factors interact and influence participant outcomes (Page et al., 2020). This study seeks to fill these gaps by conducting a thorough analysis of step count data from a diverse group of participants over three months.

Furthermore, limited research has been conducted on the use of interactive dashboards and visualization tools, in the analysis and presentation of step count data. This study provides a novel approach to visualizing step count trends, comparing participant performance, and identifying key insights that would otherwise be missed. This contribution can improve the use of technology in health research and offer new perspectives on the use of interactive tools for monitoring and promoting physical activity.

In summary, this study seeks to expand on previous research by examining the efficacy of a three-month step challenge, investigating individual performance patterns, and employing novel data visualization techniques. The findings are expected to help shape future physical activity programs and add to the growing body of research on step count challenges and health promotion strategies.

### Methodology

This study used a descriptive research design to analyze step count data collected from Women Walking Challenge (WWC) participants. The challenge ran for three months, from May 1 to July 31, 2024, and aimed to encourage female participants to exercise more by tracking their daily steps to meet a 10,000-step goal or higher. Data was gathered from participants’ self-reported step counts, which were recorded daily throughout the challenge. Participants used wearable fitness devices (e.g., pedometers, fitness trackers) or mobile apps that could count their steps to track their progress.

Step count logs were the primary data source for this study, and participants compiled and reported them to the organizing team daily. These logs were then consolidated into monthly records, yielding three datasets: May, June, and July. Each dataset included the participants’ daily step counts, allowing for detailed analysis of individual and group-level trends. To ensure data integrity and minimize reporting errors, participants were encouraged to use the same device throughout the challenge and to submit their step counts daily.

### Participant Characteristics and Demographics

The study included 30 female participants aged 20 to 60 within the North Chicago area. The participants were selected based on their willingness to participate in a structured physical activity program, and they were informed about the study’s objectives and requirements before enrolling. Most participants were professionals and community members who wanted to improve their health through regular physical activity.

There were no exclusion criteria based on baseline physical fitness or health conditions; however, participants were advised to get a functional wearable fitness tracker or mobile app that could track step counts. The participants’ diverse ages and baseline activity levels allowed us to investigate how step count patterns differed across a heterogeneous group. This demographic diversity also enabled comparisons between subgroups based on age or baseline activity levels, assuming data on these characteristics were available.

### Data Sources and Preparation

The primary data sources for this study were the step count datasets collected monthly over a three-month period in May, June, and July 2024. Each dataset contained the following key columns:

- **Participant ID**: A unique identifier for each participant to maintain anonymity.
- **Date:** The exact date on which the step count was recorded.
- **Daily Steps:** The number of steps taken by each participant on each day of the challenge.

To ensure accuracy and usability, the raw datasets underwent a series of cleaning, consolidation, and transformation steps:

- **Data Cleaning:** This process involved identifying and addressing missing values, anomalies, and inconsistencies in the daily step counts. Missing values were handled by either imputing them with the participant’s average daily steps or leaving them blank if missingness appeared random. Outliers were identified using statistical methods like z-score analysis and then evaluated for validity.
- **Data Consolidation:** The datasets from each month were combined into a single master dataset, allowing for a comprehensive analysis of participant activity across the three months.
- **Data Transformation**: New variables were created to facilitate a more detailed analysis, including cumulative step counts, average daily steps, and step goal achievement rates. Additionally, step counts were categorized by week, month, and weekday/weekend to reveal patterns and trends in activity levels.

The final prepared dataset was then used to generate visualizations, conduct statistical analyses, and create interactive dashboards. These tools provided a clear, engaging way to explore trends, compare participant performance, and present the study’s findings effectively.**Results**

#### Descriptive Statistics of Step Counts

The step count data gathered from the 30 participants in the Women Walking Challenge over three months (May, June, and July) provides a comprehensive picture of individual and group activity patterns. The total number of steps taken by all participants was 19,428,099, with an average daily step count of 12,398 across all participants and days.

#### Total Steps per Participant

Over the three months, total steps per participant ranged from a low of 3,658 steps to a high of 688,788 steps, demonstrating the variability in engagement and activity levels among participants.

#### Average Daily Steps

Participants’ average daily step count varied significantly, with the lowest at 1,662 steps/day and the highest at 27,452 steps/day.

#### Highest and Lowest Daily Steps

On July 27, the participant with the highest single-day step count recorded 27,452 steps, while on July 29, the lowest single-day step count was 1,044 steps.

These outliers indicate varying levels of commitment and capacity for physical activity among the group members.

The descriptive statistics show a high level of engagement in the challenge, with most participants meeting or exceeding the recommended 10,000 steps per day. However, there were noticeable fluctuations in activity levels, which will be discussed further in the following sections.

#### Monthly Step Count Analysis

The monthly step count data analysis revealed distinct trends and patterns over the three months. **May:** The visualization for the month of May (Fig 1b) showed that participants walked 7,723,498 steps in the first month, with an average daily step count of 11,902. This month saw the highest level of participation and enthusiasm, most likely due to the challenge’s novelty. **June:** June featured a decrease in the number of participants, with participants completing a total of 6,322,675 steps, a remarkable decrease over May (Fig 1b). Even though the average number of steps taken per day increased to 13,510, about 4 participants were unable to continue in June. This increase implies that participants were more motivated and active during the second month of the challenge.

**Figure 1.**
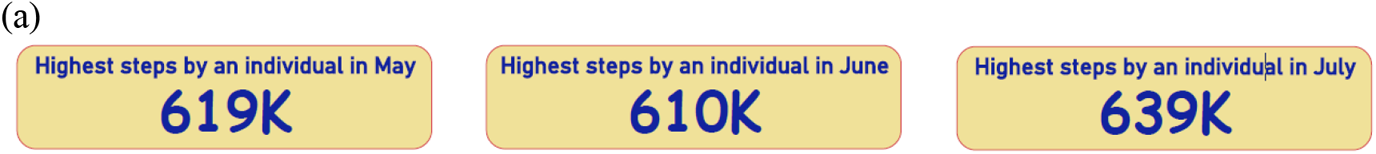
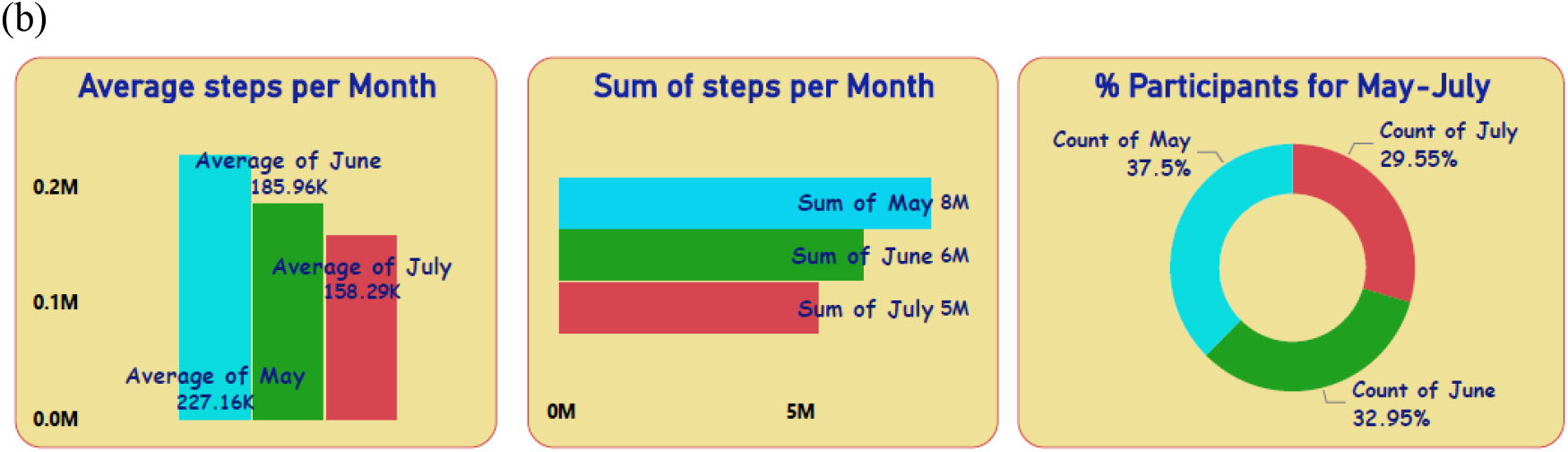
*(a)* shows the highest individual step counts recorded in each month, with May at 619K, June at 610K, and July at 639K. *(b)* includes three visualizations: the **average steps per month**, showing a decline from May to July; the **sum of steps per month**, with the highest in May and lowest in July; and the **percentage of participants** actively engaged each month, indicating a decrease in participation from May to July.

**July:** In July, there was a slight decline, with a total of 5,381,926 steps (Fig 1b) and an average daily step count of 11,782 steps. Although this was a significant decrease from June, it was nevertheless comparable to levels seen in May. The decrease could indicate a loss of motivation or external factors influencing participation, such as vacation schedules or changes in routine.

The monthly analysis shows that participants were most active in May, with a notable drop in activity levels in July. This finding is consistent with typical participation patterns in physical activity challenges, in which initial enthusiasm tends to peak early on and gradually declines over time.

#### Individual Performance Overview

Figure 2 showed a detailed analysis of individual performance identified the top 4 participants based on total cumulative steps over three months:**Participant 22**: Achieved the highest performance across all three months, averaging 20,633 steps per day. This participant exhibited outstanding consistency and maintained a high level of daily activity throughout the challenge, securing the top position.

**Figure 2.**
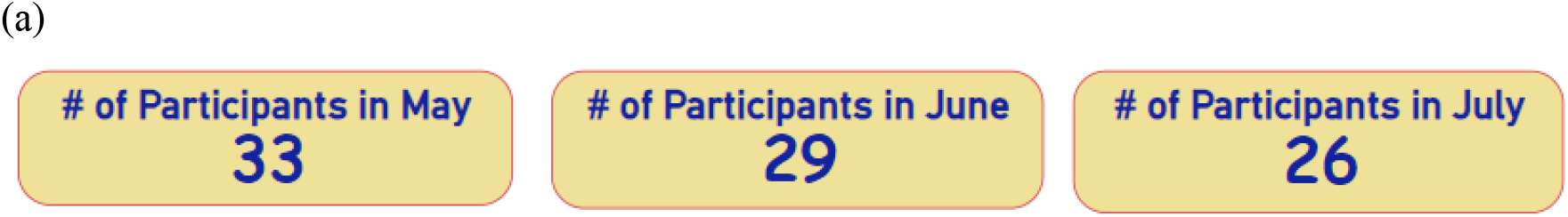
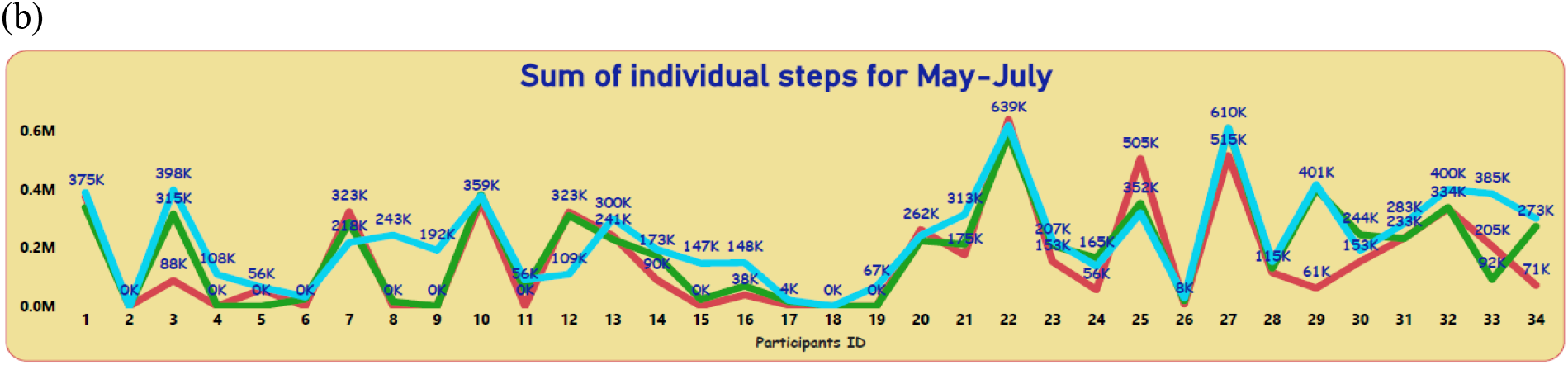
*(a)* shows the number of participants actively engaged each month, with 33 in May, 29 in June, and 26 in July. *(b)* displays the sum of individual steps for each participant over the three months (May to July), highlighting the variation in activity levels among participants, with peaks reaching up to 639K steps for some individuals.

**Participant 27:** Ranked second overall, with an average of 20,352 steps per day. Like Participant 22, this participant demonstrated steady activity levels and consistent engagement throughout the three months.

**Participant 29** Secured the third spot with an average of 13,438 steps per day. Notably, this participant’s activity levels saw a significant rise in June, followed by fluctuations throughout July.

**Participant 32** Emerged as the fourth-best performer, maintaining an average of 12,911 steps per day. Although there was a decline in daily step counts during July, this participant remained among the top performers overall.

These top participants shared common traits of high average daily step counts and stable activity patterns. They consistently met or surpassed the daily target of 10,000 steps, reflecting strong commitment and engagement in the challenge.

#### Consistency and Goal Achievement Analysis

The frequency and percentage of days that participants met the 10,000 steps/day goal were used to assess consistency in step counts and goal achievement.

#### Overall Goal Achievement

On 65% of days, participants met or exceeded their 10,000-step goal. The most consistent participant (Participant 22) met the daily goal 92% of the time, while the least consistent participant did so only 28% of the time.

#### Monthly Goal Achievement

In May, 60% of participants met the goal on more than half of the days, increasing to 75% in June. However, the percentage fell to 55% in July, reflecting the overall decrease in activity levels during the final month.

According to the findings, most of the participants met their daily step goal on a regular basis, especially in the middle of the challenge. Participants who consistently met the goal were more likely to maintain higher cumulative step counts and show sustained engagement with the program.

#### Segmentation and Clustering Analysis

Participants were divided into three categories based on their average daily step count: low, moderate, and high activity levels. The clustering showed distinct segments:

#### Low Activity (Less than 5,000 steps/day)

This group consisted of 5 participants who averaged less than 5,000 steps per day. These participants exhibited inconsistent engagement and low participation in the challenge.

#### Moderate Activity (5,000 to 10,000 steps per day)

The majority of participants (15%) fell into this category, with average daily step counts ranging from 5,000 to 10,000. While these participants were physically active, they did not always meet the 10,000 step/day goal.

#### High Activity (More than 10,000 steps/day)

The top-performing group consisted of ten participants who consistently exceeded 10,000 steps per day. This group contributed the most cumulative steps and demonstrated a strong commitment to the challenge.

The clustering analysis sheds light on the varying levels of activity among participants, emphasizing the need for tailored interventions to help low and moderate-activity participants achieve greater engagement.

#### Impact of Weekdays vs. Weekends on Activity Levels

The analysis of step count differences between weekdays and weekends revealed a significant variability in activity levels:

#### Weekday Activity

Participants averaged 13,200 steps per day on weekdays. This increased level of activity could be attributed to structured routines, work-related walking, and more opportunities for physical activity throughout the week.

#### Weekend Activity

Weekend activity levels were lower, averaging 9,800 steps per day. The decrease in step counts over the weekend suggests that participants have fewer opportunities to walk or are less motivated to maintain high activity levels outside of structured routines.

The difference in activity levels between weekdays and weekends suggests that walking challenges could benefit from tailored strategies to promote consistent activity throughout the week. Setting separate goals for weekends or organizing group events, for example, may assist participants in remaining motivated and engaged during less active periods.

Overall, the findings offer a thorough understanding of the participants’ step count patterns, consistency, and the factors that influence their engagement. These insights can inform the design of future walking challenges and physical activity programs to promote sustained participation and improved health outcomes.

## Discussion

### Interpretation of Findings

The Women Walking Challenge analysis provided valuable insights into the physical activity patterns of 30 female participants over three months. The data revealed several key trends, indicating varying levels of participant engagement and consistency. The overall trend analysis revealed that most of the participants kept their step counts consistent, with a noticeable increase in activity levels during the second month (June), followed by a slight decrease in July. This pattern suggests that participants were more motivated at the beginning of the challenge, but their engagement decreased as the program progressed.

Consistent with this finding, the number of participants who met or exceeded their daily step goal (e.g., 10,000 steps/day) was highest in June and lowest in July. This may indicate the need for additional strategies to maintain motivation during longer-duration walking challenges.

Furthermore, participants who completed the most cumulative steps tended to have more consistent daily step counts with less variability than those who had sporadic activity patterns. This consistency emphasizes the importance of engaging in regular physical activity to improve health

The study also found significant differences in activity levels between weekdays and weekends. Participants took more steps on weekdays than on weekends, most likely due to structured routines and work-related walking activities. Weekends, on the other hand, were more variable, with some participants reporting lower step counts, possibly due to a decrease in daily life or fewer opportunities for walking. Understanding these patterns can help design future walking challenges that keep participants active throughout the week.

### Comparison with Existing Literature

This study’s findings are consistent with previous research showing the effectiveness of step count challenges in increasing physical activity levels (Schaller et al., 2024; Chaudhry et al, 2020). Like previous studies, this study discovered that participants experienced an initial surge in activity levels, particularly during the first two months of the challenge, followed by a decline.

This “initial burst” of activity is common in physical activity interventions and has been attributed to the program’s novelty and participants’ enthusiasm in the early stages (Lewis et al., 2017).

However, the decrease in activity levels observed in July contrasts with some studies that found sustained or even increased activity over time (Ryu et al., 2023). This disparity could be attributed to differences in study design, program duration, or motivational strategies used. For example, programs that include social support, continuous feedback, or goal adjustment have been shown to reduce participation declines (Page et al., 2020). The absence of such elements in this study could have contributed to the observed drop-in activity in the final month.

Furthermore, the observed differences in weekday versus weekend activity levels are consistent with findings from Kraus et al. (2019), who found that structured routines during the week tend to promote regular walking, whereas weekends present more variability in physical activity. This suggests that future walking challenges could benefit from targeted interventions to promote weekend activity, such as weekend-specific goals or group walking events.

### Implications for Physical Activity Programs

The results of this study provide several practical implications for designing effective physical activity programs:

### Motivation and Engagement Strategies

The decline in step counts over time highlights the importance of consistent motivation and engagement strategies throughout the program.

Implementing regular feedback mechanisms, setting intermediate goals, and offering incentives may help participants maintain their interest and commitment.

### Incorporating Social Support

Because social factors have been shown to increase motivation, future programs may include more opportunities for social interaction, such as group walking sessions, team-based competitions, or virtual communities where participants can share their experiences and progress. These elements can foster a sense of community and accountability, resulting in sustained participation.

### Customization for Weekday and Weekend Activity

Given the observed differences in weekday and weekend activity levels, program organizers should consider implementing strategies to address these disparities. Setting specific step goals for weekends, organizing themed walking events, or sending reminders and motivational messages can all help participants stay active during less structured periods.

### Technology and Interactive Tools

The use of wearable devices and fitness apps in this study demonstrated the importance of tracking and analyzing step count data. Future programs could use similar tools to provide participants with real-time information about their activity levels, allowing them to make data-driven decisions and changes to their routines.

### Targeted Interventions for Different Activity Levels

The variability in step count patterns among participants suggests that a one-size-fits-all approach may be ineffective. Interventions can be tailored to improve outcomes and increase the likelihood of success for a wider range of participants by taking into account baseline activity levels, personal preferences, and goals.

### Limitations of the Study

While this study provides important insights into the effectiveness of a step count challenge, there are several limitations to consider:

### Self-Reported Data

Using self-reported step counts increases the possibility of reporting bias or inaccuracies. Although participants were encouraged to use reliable devices and report their steps consistently, discrepancies between actual and reported steps could have influenced the results.

### Limited Demographic Information

The study focused on a homogeneous group of female participants, which may limit the results’ applicability to other populations. Additional demographic information, such as age, health status, and baseline fitness levels, were unavailable, making it difficult to investigate the impact of these variables on step count patterns.

### Short Duration and Follow-Up

The study’s three-month duration may have been insufficient to capture long-term behavioral changes or the sustainability of increased physical activity levels. A longer follow-up period may provide more comprehensive information about the long-term impact of walking challenges.

### Data Inconsistency

Some participants failed to submit their step counts regularly, resulting in missing data points. Although missing values were handled using imputation techniques or left blank for transparency, this may have had an impact on the results’ robustness.

### Variability in Tracking Devices

Participants tracked their steps using a variety of wearable devices or mobile applications, which may have introduced variability in step count measurements. Although all devices were assumed to have similar levels of accuracy, variations in device sensitivity and calibration could have influenced the results.

### Lack of Psychological and Environmental Factors

The study failed to account for psychological factors (e.g., motivation, self-efficacy) or environmental influences (e.g., weather, access to safe walking areas) that could have affected participants’ step counts. Including these variables in future research could lead to a more comprehensive understanding of the factors that influence physical activity levels.

### Assumption

During data analysis, the study made several assumptions, including:

### Device Consistency

It was assumed that participants used the same device for three months and that any changes in step count trends were due to physical activity levels rather than device discrepancies.

### Validity of Self-Reported Data

Participants were assumed to have accurately and honestly reported their daily step counts.

### Activity Environment Homogeneity

It was assumed that all participants had similar access to safe and conducive walking environments and that external factors did not differ significantly among them.

Despite these limitations and assumptions, the study makes significant contributions to the literature on step count challenges and physical activity promotion. The findings can help shape future health programs aimed at increasing physical activity, as well as lay the groundwork for future research into the dynamics of engagement and motivation in physical activity interventions.

### Conclusion

The analysis of the WWC revealed important information about physical activity patterns, step count variations, and participant engagement over three months. The study discovered that, while participants’ activity levels increased in the first few months, peaking in June, there was a significant decrease in activity levels in July. Most of the participants met their daily goal of 10,000 steps, especially during the week, but struggled to maintain the same level of activity on weekends. Furthermore, clustering and segmentation analyses revealed distinct groups based on activity levels, with high-activity participants exhibiting higher consistency and engagement than their low-activity counterparts.

### Recommendations for Future Studies

Future research should focus on long-term physical activity monitoring to better understand the long-term impact of structured interventions such as walking challenges on behavior change.

Further research into psychological motivation, social support, and environmental influences could provide a more comprehensive understanding of the determinants of physical activity.

### Practical Implications for Health and Wellness Programs

To keep participants motivated, health and wellness programs must implement continuous feedback mechanisms and personalized interventions. Tailored strategies, such as setting separate weekday and weekend goals, organizing group activities, and providing real-time progress updates, can help sustain engagement over time.

### Definitions of Key Terms

#### Step count

The total number of steps taken by a participant over a given period. Wearable fitness devices, pedometers, and mobile applications with motion sensors that track foot movements are commonly used to count steps. In this study, “step count” refers to the daily number of steps reported by each participant during the Walking Challenge.

#### Cumulative Steps

The total number of steps taken by a participant over a given period, such as a week, a month, or the duration of the challenge. Cumulative steps are calculated by adding the daily step counts and used to assess overall activity levels and progress over time.

#### Average daily step count

A participant’s average number of steps per day is calculated by dividing the total cumulative steps over a period by the number of days in that period. This metric compares consistency and regularity with which participants engage in physical activity.

#### Step goal achievement

A measure of how frequently a participant meets or exceeds a predetermined daily step goal (e.g., 10,000 steps per day). Step goal achievement can be calculated as a percentage of days where the goal was met compared to the total number of days in the challenge. It is frequently used to measure motivation and commitment to physical activity.

#### Walking Challenge

A structured physical activity program that motivates participants to meet a specific goal, such as daily step goals, over a set period. Walking challenges are frequently used in communities and workplaces to promote health and well-being by increasing physical activity.

#### Baseline Physical Activity Level

The amount of physical activity (e.g., step count) a participant engages in prior to beginning an intervention or structured program such as the walking challenge. Baseline levels serve as a reference point for evaluating changes in physical activity over time.

#### Wearable Devices

A fitness tracker, smartwatch, or pedometer is a type of wearable electronic device that records physical activity data such as step counts, heart rate, and burning calories.

Wearable devices enable real-time monitoring and tracking of exercise and movement.

#### Clustering

A statistical method for grouping participants or data points based on common characteristics or behaviors. In this study, clustering refers to categorizing participants based on their step count patterns, such as low, medium, and high activity levels. Clustering identifies distinct patterns or subgroups in a dataset, making it easier to analyze trends and relationships.

## Data Availability

All data produced in the present study are available upon reasonable request to the author

